# Involvement of the habenula in the pathophysiology of autism spectrum disorder

**DOI:** 10.1101/2021.04.08.21255122

**Authors:** Jürgen Germann, Flavia Venetucci Gouveia, Helena Brentani, Saashi A. Bedford, Stephanie Tullo, M. Mallar Chakravarty, Gabriel A. Devenyi

## Abstract

The habenula is a small epithalamic structure that has rich widespread connections to multiple cortical, subcortical and brainstem regions. It has been identified as the central structure modulating the reward value of social interactions, behavioral adaptation, sensory integration and circadian rhythm. Autism spectrum disorder is characterized by social communication deficits, restricted interests and repetitive behaviors, and frequently associated with altered sensory perception and mood and sleep disorders. The habenula is implicated in all these behaviors and results of preclinical studies suggest a possible involvement of the habenula in the pathophysiology of this disorder. Using anatomical magnetic resonance imaging and automated segmentation we show that the habenula is significantly enlarged in children and adults with ASD compared to age matched controls. The present study is first to provide evidence of the involvement of the Hb in the pathophysiology of ASD.

## Introduction

The habenula (Hb) is a small phylogenetically preserved epithalamic structure that plays a key role in the control of the monoaminergic system ^1,2^. It is divided into lateral and medial parts based on characteristic cytoarchitectonic and connectivity patterns. Through its rich widespread connections, especially to the hypothalamus, limbic areas and brainstem nuclei (illustrated ^3^ in Figure 1) the habenula is implicated, among others, in social interaction, reward processing, behavioral adaptation, sensory integration and circadian rhythm (Figure 2) ^1,2,4–9^.

**Figure 1:**
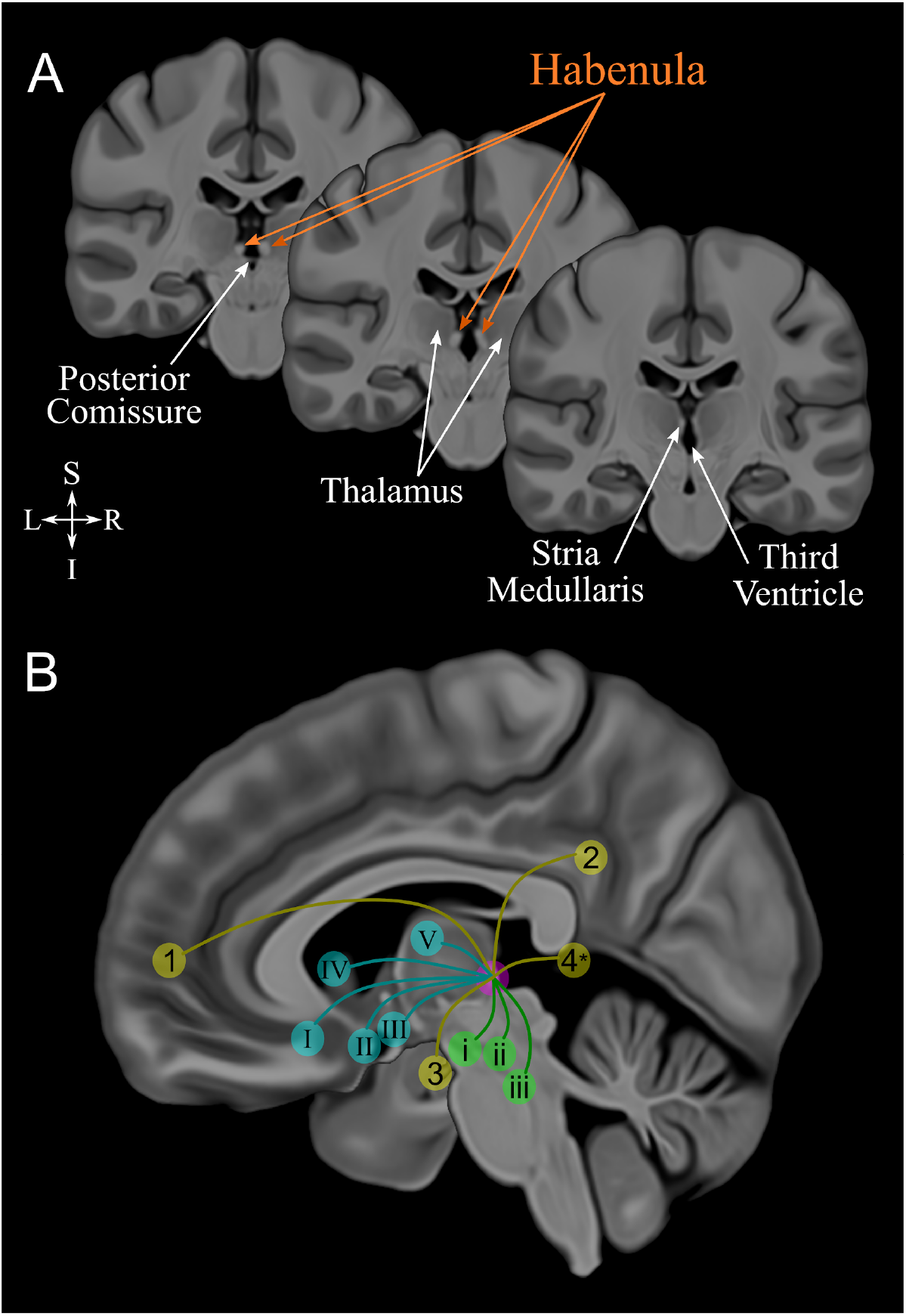
Habenula anatomy, boundaries and connections displayed using a high-resolution, high contrast template by Neudorfer and colleagues ^3^. **A**. Coronal slices illustrating the location of the Habenula, a structure appearing bright (hyperintense) on T1 weighted magnetic resonance images, surrounding structures and its boundaries. **B**. Diagram illustrating the connectivity of the Hb. Cortical regions cortical in yellow: 1, medial prefrontal cortex; 2, cingulate gyrus; 3, hippocampus & parahippocampal gyrus; 4, posterior insula (*estimated location). Subcortical regions in blue: I, basal forebrain; II, hypothalamus; III, nucleus basalis of Meynert; IV, basal ganglia; V, thalamus. Brainstem regions in green: i, ventral tegmental area; ii, substantia nigra; iii, periaqueductal grey - raphe nuclei.

**Figure 2:**
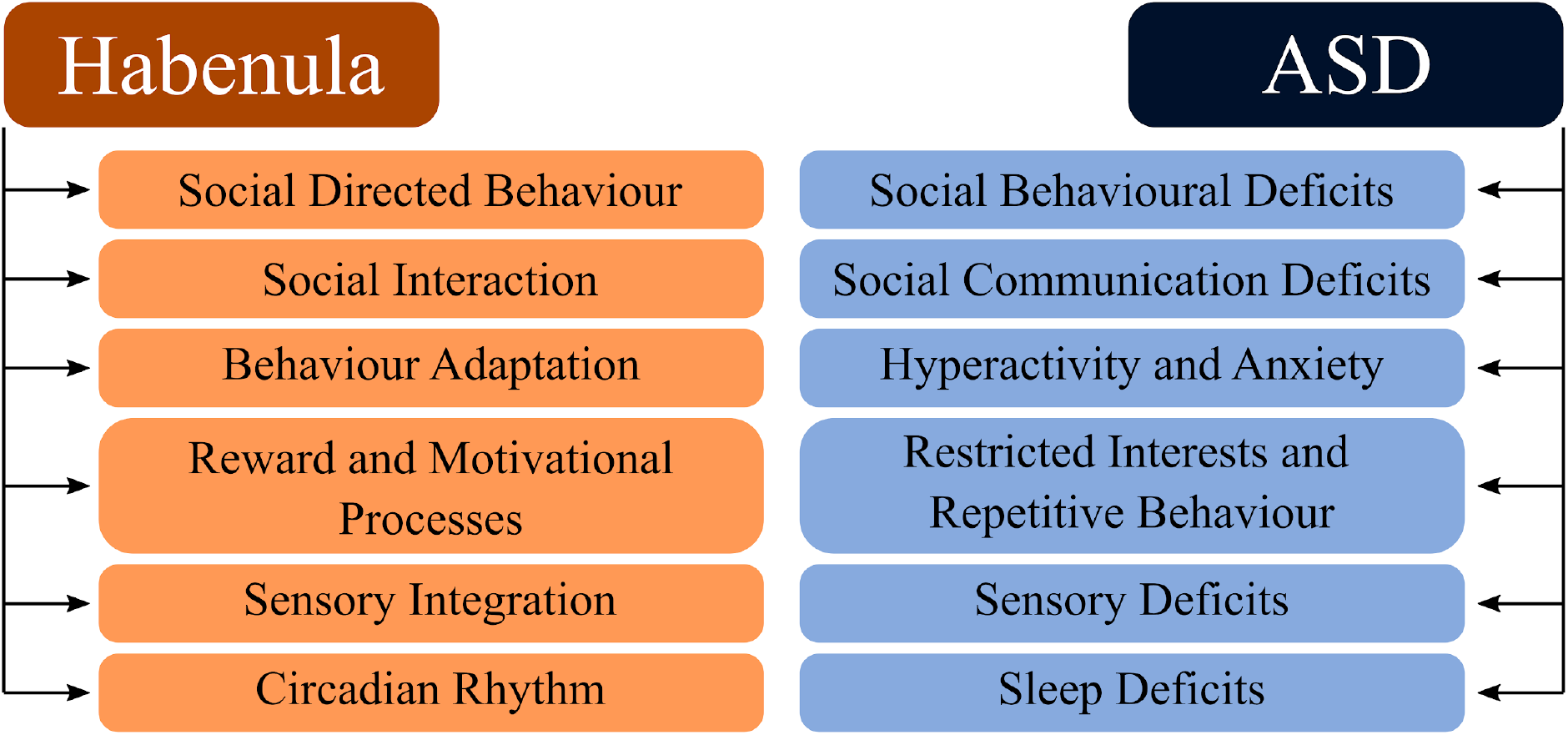
Functions that the Habenula is critically involved in and deficits found in autism spectrum disorder. Abbreviation: ASD: autism spectrum disorder.

Most of the research investigating the role of the Hb in social behaviour has been performed in preclinical settings and while neuroimaging and brain stimulation techniques have provided some insight into the role of the Hb in psychiatric disorders further research is warranted to obtain a deeper understanding of the neurocircuitry of social behaviour and relate it to the deficits found in human disorders ^10,11^. Autism Spectrum Disorder (ASD) is classically characterized by social communication deficits, restricted interests and repetitive behaviors, and frequently associated with altered sensory perception and mood and sleep disorders (Figure 2) ^12,13^. Thus, an involvement of the Hb in the neurobiology of ASD is plausible but has yet to be demonstrated in humans. Here, we investigated the hypothesis that the Hb plays a role in ASD by analyzing morphometric Hb characteristics in a large cohort of ASD subjects (218 subjects; 182 males) and age matched Healthy Controls (HC; 302 subjects; 212 males) from the Autism Brain Imaging Data Exchange (ABIDE) repository ^14,15^, with an age range spanning from early childhood to adulthood (6 to 30 years).

## Results

The morphometry (volume) of the left and right habenula of each subject was obtained through automatic segmentation (Figure 3A) ^16,17^. Comparing total bilateral Hb volume (co-varied withage, sex and total brain volume with the study site used as a random intercept effect) we found that ASD subjects have significantly larger bilateral Hb volumes compared to HC (ASD subjects: 27.1 mm^3^±5.3; HC: 25.5 mm^3^±4.5; t=3.28, p=0.001; Figure 3B). This significant volume difference is apparent across the entire age range studied (6 to 30 years; Figure 3C, see Figure 3D for 3-D reconstruction of the habenula) unlike what has previously been reported in Schizophrenia and Bipolar disorder where there is a significant habenula volume difference between patients and controls only in adults and not in children or adolescents

**Figure 3:**
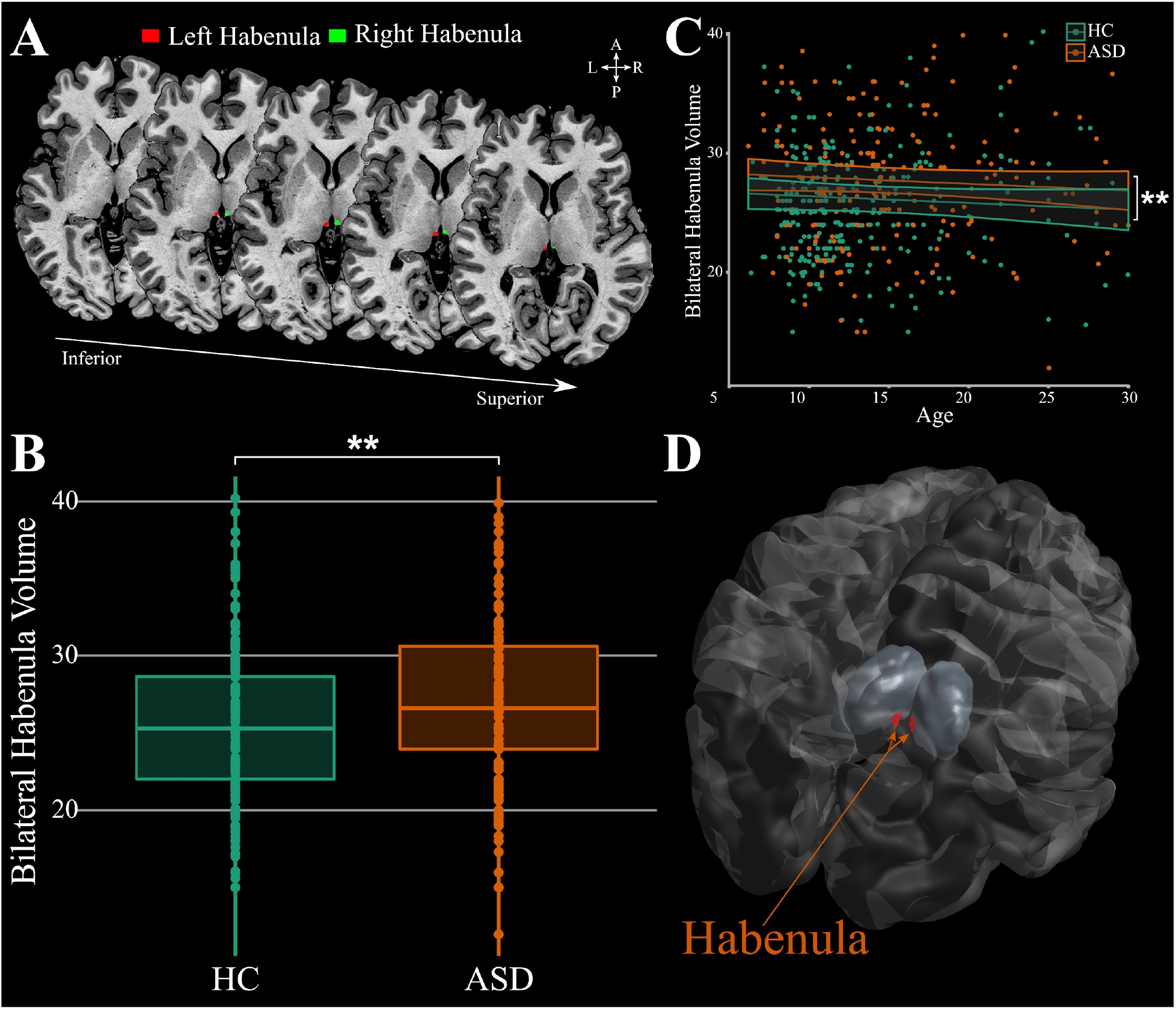
Habenula volume differences found in ASD. **A**. Example MAGeTBrain habenula segmentation on a T1 weighted magnetic resonance image illustrated on the axial plane. **B-C**. Bilateral habenula volume is greater in ASD compared to HC and this effect is independent of age. **D**. 3D reconstruction of the habenula. Bilateral habenula volume was corrected for age, sex and total brain volume. ** indicates p≤0.01. Abbreviations: ASD: subjects with autism spectrum disorder; HC: healthy control subjects.

## Discussion

While T1w contrast does not allow one to discern the underlying cause of these volume differences (e.g. increased dendrites, microglia, angiogenesis, neuroglia) previous work in animals showed that increased Hb activity is associated with social deficits that are ameliorated when Hb activity is inhibited ^6^. The Hb has been identified as the central structure mediating the reward value of social interactions and shown to be a key region for adapting behavioural strategies, integrating both internal (previous reward experience) and external (sensory input) information to initiate necessary behavioural adjustments ^2,5^. Furthermore, an optimal Hb function is necessary for flexible behavioural adjustments ^2,5^, sensory processing ^2,7^, motivational processes ^1,2^ and regulation of circadian rhythms ^4^ all domains where deficits are found in ASD ^12,13^ (e.g. social communication deficits, restricted interests and repetitive behaviors, altered sensory perception and mood and sleep disorders; Figure 2).

The increase in Hb volume demonstrated in this study strongly suggests that the Hb is at the core of the neurocircuitry of ASD potentially involved in a broader range of behavioral symptoms, beyond the classic deficits of social behaviour and social interaction. Further studies investigating neurotransmitters, metabolites, connectivity patterns and neurochemical binding are necessary to unravel the neurobiological mechanisms underlying the involvement of the Hb in ASD. While previous studies using preclinical models have demonstrated the important role of the Hb for social behaviour and social interaction ^6,9^ the present study is the first to document an increased volume of the Hb in ASD patients, pointing to the involvement of the Hb in the pathophysiology of ASD.

## Materials and Methods

*Subjects:* Habenula (Hb) volumes were estimated using T1-weighted magnetic resonance images (MRI) scans of subjects from the Autism Brain Imaging Data Exchange (ABIDE) repository ^14,15^. ABIDE is a large-scale, publicly available multi-site database providing MRI scans as well as behavioral data from individuals with autism spectrum disorder (ASD) and healthy controls (HC). For detailed demographics, imaging acquisition parameters, local institutional review boards and written informed consent, please visit (http://fcon_1000.projects.nitrc.org/indi/abide/). Inclusion criteria were motion and scan quality as previously described ^18^, complete Social Responsiveness Scale scores ^19^, sufficient data to model the developmental trajectories in ASD subjects and HC. This resulted in a final dataset of 218 ASD subjects (182 males) and 302 HC (212 males) ages 6 to 30 years.

### Image Processing and MAGeTbrain Segmentation

The images were pre-processed using iterative non-uniformity correction and skull-stripped (https://github.com/CoBrALab/minc-bpipe-library). The Multiple Automatically Generated Templates (MAGeT) brain segmentation algorithm was used to segment the Hb (https://github.com/CoBrALab/MAGeTbrain) ^16,17^. MAGeTbrain employs label propagation to produce individual segmentations using five segmented high-resolution atlases. These atlases are then propagated to 21 template images selected from the input dataset. In doing so a large number (5×21=105) of candidate segmentations is created, which are fused using majority vote to derive a final segmentation for each subject. Using this template library has two advantages: it helps reduce atlas bias and it reduces registration errors by averaging ^20^. To ensure accurate segmentation, the resulting individual Hb segmentation of each subject was visually inspected by two raters (JG and FVG) in 3D overlaid onto the individual T1-weighted MRI image using DISPLAY (https://www.mcgill.ca/bic/software/minc/minctoolkit).

### Statistical analysis

The lme4 (version 1.1-21) and lmerTest (version 3.1.1) packages in R (version 3.6.1; https://www.r-project.org) were used to perform the statistical analysis. Bilateral Hb volumes (left Hb + right Hb) were calculated and used for subsequent analysis. A linear mixed effect model corrected for age, sex and total brain volume with the site used as a random intercept effect was used to test for a possible effect of diagnosis on individual Hb volume.

## Data Availability

The dataset analysed during the current study (Autism Brain Imaging Data Exchange, ABIDE I & ABIDE II) is publicly available at http://fcon_1000.projects.nitrc.org/indi/abide.

The tool used in this study for automatic segmentation of the habenula is freely available at https://github.com/CoBrALab/MAGeTbrain. The processed data are available from the corresponding author upon reasonable request.

## Acknowledgements

GAD acknowledges that this work was supported in part by funding provided by Brain Canada, in partnership with Health Canada, for the Canadian Open Neuroscience Platform initiative. This research was enabled in part by support provided by SciNet (www.scinethpc.ca) and Compute Canada (www.computecanada.ca).

## Author Contributions

JG analyzed and interpreted the data and contributed to all drafts of the manuscript; FVG wrote the draft manuscript and its revisions; SAB, ST and MMC performed data quality control and contributed to all drafts of the manuscript; HB and GAD contributed to all drafts of the manuscript.

## Data Availability Statement

The dataset analysed during the current study (Autism Brain Imaging Data Exchange, ABIDE I & ABIDE II) is publicly available at http://fcon_1000.projects.nitrc.org/indi/abide/. The tool used in this study for automatic segmentation of the habenula is freely available at https://github.com/CoBrALab/MAGeTbrain. The processed data are available from the corresponding author upon reasonable request.

